# Germline prediction of immune checkpoint inhibitor discontinuation for immune-related adverse events

**DOI:** 10.1101/2024.06.10.24308518

**Authors:** Pooja Middha, Rohit Thummalapalli, Zoe Quandt, Karmugi Balaratnam, Eduardo Cardenas, Christina J. Falcon, Princess Margaret Lung Group, Matthew A. Gubens, Scott Huntsman, Khaleeq Khan, Min Li, Christine M. Lovly, Devalben Patel, Luna Jia Zhan, Geoffrey Liu, Melinda C. Aldrich, Adam J. Schoenfeld, Elad Ziv

## Abstract

**Introduction:** Immune checkpoint inhibitors (ICIs) can yield remarkable clinical responses in subsets of patients with solid tumors but can also often lead to immune-related adverse events (irAEs). Predictive features of clinically severe irAEs leading to cessation of ICIs have yet to be established. Using data from 1,327 patients with lung cancer treated with ICIs between 2009 and 2022 at four academic medical centers, we evaluated the association of a germline polygenic risk score for autoimmune disease and discontinuation of ICIs due to irAEs.

**Methods:** Using Cox proportional hazards model, we assessed the association between a polygenic risk score for autoimmune disease (PRS_AD_) and cessation of ICI therapy due to irAEs. All models were adjusted for age at diagnosis, sex, lung cancer histology, type of therapy, recruiting center, and the first 5 principal components. To further understand the differential effects of type of therapy and disease stage on the association between PRS_AD_ and cessation of ICI due to irAEs, we conducted stratified logistic regression analysis by type of ICI therapy and disease stage.

**Results:** We found an association between PRS_AD_ and ICI cessation due to irAEs (HR per SD = 1.18, 95% CI = 1.02 – 1.37, P = 0.03). This association was particularly strong in patients who had ICI cessation due to irAEs within three months of therapy initiation (HR per SD = 1.38, 95% CI = 1.08 - 1.78, P = 0.01). Individuals in the top 20th percentile of PRS_AD_ had 7.2% ICI discontinuation for irAEs by three months, compared to 3.9% discontinuation by three months among patients in the bottom 80th percentile (log-rank P = 0.02). In addition, among patients who received combination PD-1/PD-L1 and CTLA-4 inhibitor therapy, PRS_AD_ had an OR per SD of 1.86 (95% CI = 1.08 - 3.51, P = 0.04).

**Conclusions:** We demonstrate an association between a polygenic risk score for autoimmune disease and early ICI discontinuation for irAEs, particularly among patients treated with combination ICI therapy. Our results suggest that germline genetics may be used as an adjunctive tool for risk stratification around ICI clinical decision-making in solid tumor oncology.

## Introduction

Immune checkpoint inhibitors (ICIs), including anti–PD-1/PD-L1 and anti-CTLA-4 based therapies^1–3^, have revolutionized treatment landscapes across a wide spectrum of solid tumors including cutaneous^4,5^, lung^6,7^, genitourinary^8,9^, hepatobiliary^10^, and many other malignancies, and can lead to durable clinical responses in subsets of patients. However, in the clinic, the predicted benefit of ICIs must be balanced by the risk of incumbent immune-related adverse events (irAEs) resulting from enhanced immune system activation, with irAEs of any grade estimated to occur in approximately 30-40% of patients treated with ICIs across indications^11–13^ Although irAEs are often mild to moderate in severity, the incidence of high-grade irAEs has been estimated to be up to 8-20% for anti-PD-1/PD-L1 monotherapy^14–16^ and up to 18-59% for combination anti-PD-1 and anti-CTLA-4 therapy^4,16^. In some cases, irAEs can lead to severe morbidity and even patient death^17,18^, and subsets of irAEs have been shown to be irreversible^19^. The development of clinically significant irAEs can therefore often necessitate cessation of ICI therapy in the clinic.

Despite these risks, however, there remain no clear predictive features for the risk of development of clinically significant irAEs routinely considered in practice, apart from a history of prior autoimmune disease. Development of such predictive features would help optimize patient selection for ICI treatment, particularly in settings in which there remains clinical equipoise around the magnitude of predicted ICI benefit^20,21^. Efforts to characterize potential predictors of irAE development have included investigation of shared antigens in tumors and index organs experiencing irAE toxicity^22^, comprehensive gut microbiome profiling^23^, and evaluation of baseline serum autoantibodies by proteomics^24^ in individual tumor types.

Multiple recent studies have highlighted the impact of germline genetic variation on the risk of irAE development across solid tumors^25,26^, with recent work by our group describing the association of individual polygenic risk scores for hypothyroidism^27^ and ulcerative colitis^28^ with risk of development of ICI-induced thyroiditis^27^ and colitis^28^ respectively. Given the clinical value in developing a predictive marker of clinically severe irAEs more generally, we sought to understand whether germline genetics could predict the development of irAEs leading to cessation of ICI therapy. Here, using a large pooled cohort of patients with lung cancer treated with ICIs, we demonstrate the utility of a germline polygenic risk score for autoimmune disease^29^ in predicting cessation of ICIs due to irAEs.

## Methods

### Study sample

An overview of the Genetics of immune-related adverse events and Response to Immunotherapy (GeRI) study and analytical pipeline is shown in Figure 1. The GeRI study is a cohort of 1,327 patients with lung cancer who were treated with immune checkpoint inhibitors (ICIs) across four medical centers: Vanderbilt University Medical Center (VUMC), Princess Margaret Cancer Center (PM), University of California, San Francisco (UCSF), and Memorial Sloan Kettering Cancer Center (MSK). All patients were administered at least one dose of either anti-PD-1 or anti-PD-L1 monotherapy or combined with anti-CTLA-4 and/or chemotherapy.

**Figure 1:**
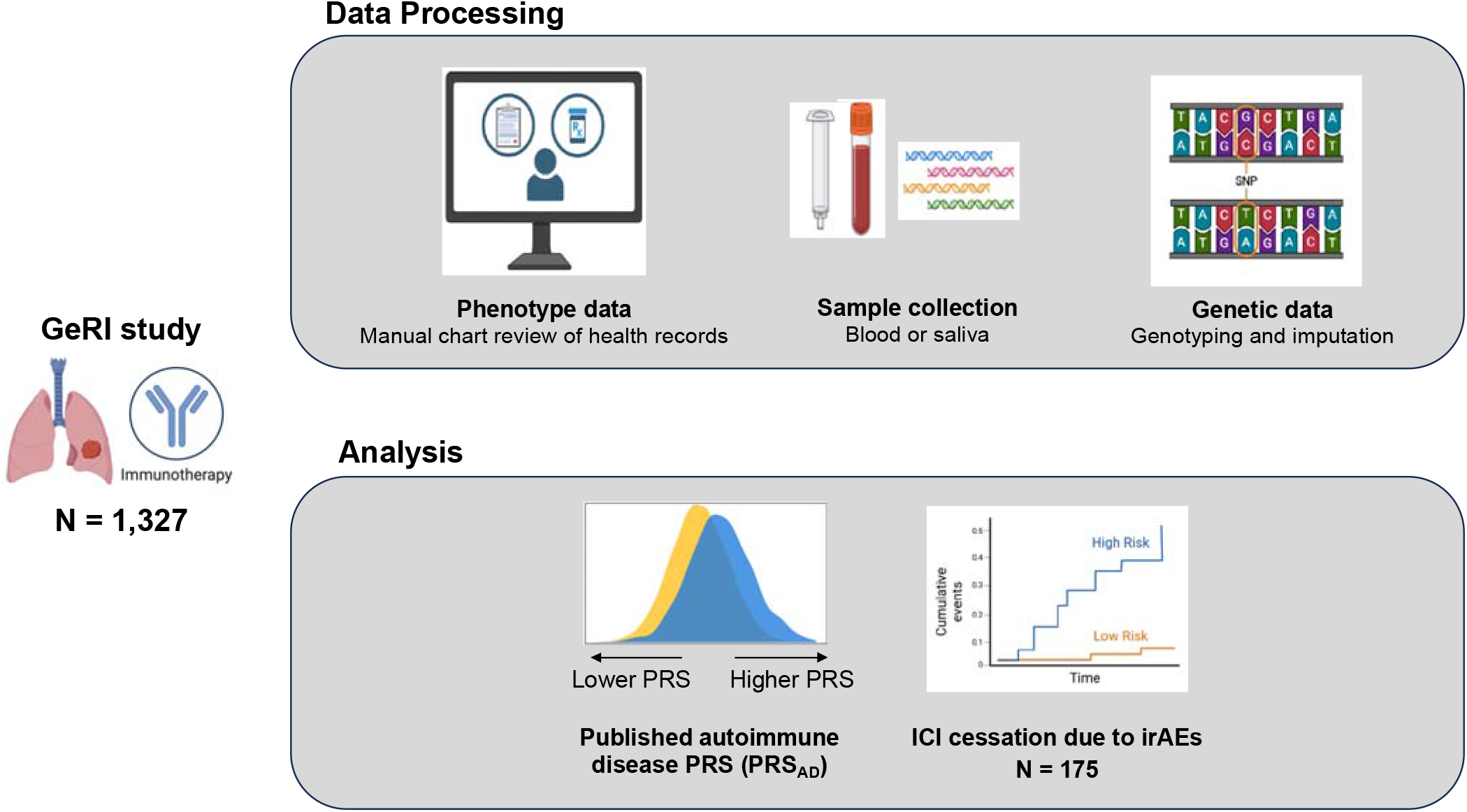
Brief overview of GeRI study and analytical pipeline. The GeRI study is comprised of 1,328 patients with non-small cell lung cancer treated with at least one dose of immune checkpoint inhibitor (ICI) therapy. Phenotype data was manually curated from health records and each participant was provided with either a blood or saliva sample for genotyping. Genotyping was performed using Affymetrix Precision Medicine Diversity Array and imputed to 1000 genomes references panel (phase 3 v5). Association analysis between a previously published polygenic risk score for autoimmune disease (PRS_AD_)^1^ and ICI cessation due to immune-related adverse events (irAEs) was conducted using the Cox proportional hazards model and Fine and Gray sub-distribution hazards models (to account for competing risks). Cumulative incidence curves were obtained by genetic risk based on PRS_AD_ percentile. Additionally, Cox proportional hazard models were performed to assess the association between PRS_AD_ and progression-free and overall survival. Kaplan-Meier survival curves and log-rank tests were performed.

Clinical and demographic data, including treatment dates and reasons for ICI therapy discontinuation, were extracted from each medical center through a manual review of medical, laboratory, and pharmacy records. The PM cohort is comprised of 306 ICI-treated patients enrolled between 2011 and 2022, while the VUMC cohort includes 265 patients who underwent ICI therapy between 2009 and 2019. VUMC participants were part of BioVU, Vanderbilt’s biomedical DNA repository linked to the de-identified health records. The MSK cohort is comprised of 677 patients treated with ICIs between 2011 and 2022. The UCSF cohort is comprised of 79 patients who received ICIs between 2019 and 2021. Local Institutional Review Boards approved the study at each center. All patients provided written informed consent.

### Genotyping and quality control

Patients from all recruiting centers provided either blood or saliva samples for genotyping. Extracted DNA was genotyped using the Affymetrix Precision Medicine Diversity array and data were imputed to 1000 genomes reference panel (phase 3 version 5) using Michigan Imputation Server. Standard quality control measures were conducted, and samples with a call rate < 95% were excluded from the analysis. Similarly, variants with genotyping rate < 95% and minor allele frequency (MAF) < 1% were excluded.

### Polygenic risk score (PRS)

We used a previously developed polygenic risk score for autoimmune disease developed by Weissbrod, et al.^29^ (PRS_AD_) to evaluate the association between PRS_AD_ and ICI discontinuation due to irAEs. Briefly, a dichotomous autoimmune disease phenotype was created by collating a spectrum of autoimmune diseases, and a cross-population PRS was developed using the PolyPred method^29^. The PRS_AD_ comprises 159,127 genetic variants with a MAF > 1%. Weissbrod et al. computed the effect estimate for each SNP using a weighted combination of tagging effect from BOLT-LMM and posterior mean causal effect estimates from functionally informed fine-mapping using PolyFun and then validated the PRS_AD_ in an independent cross-population data.

### Statistical Analysis

Using the weights from Weissbrod et al.^29^, we computed a weighted PRS_AD_ for all patients in the GeRI study. We standardized the PRS_AD_ and categorized patients into high genetic risk (> 80^th^ percentile) and low/moderate risk (≤ 80^th^ percentile). Cumulative incidence estimates by PRS_AD_ categories were computed using Kaplan-Meier curves and log-rank testing. Cause-specific hazard models were employed to assess the hazard ratio of the continuous (per standard deviation [SD]) and categorical PRS_AD_ on overall, early, and late cessation of ICIs due to irAEs. Early ICI discontinuation due to irAEs was defined as discontinuation of ICI therapy within 3 months of the start of therapy due to irAEs; late discontinuation was defined as discontinuation of ICI due to irAEs at or after 3 months of therapy. To address competing risks, such as discontinuation of ICI for reasons other than the event of interest, we performed a Fine-Gray sub-distribution hazard analysis. Additionally, we tested the PRS_AD_ for association with individual irAE subtypes using a logistic regression model. All models were adjusted for age at diagnosis, sex, lung cancer histology, type of therapy, recruiting center, and the first 5 principal components (PCs). Next, we conducted stratified analyses by type of ICI therapy and disease stage. Analyses were conducted using PLINK, R v4.2.3 (R foundation for Statistical Computing), and all P values were two-sided.

## Results

### GeRI study characteristics

We analyzed data from a total of 1,327 patients with lung cancer treated with ICIs (Table 1). In this study, the median age at lung cancer diagnosis was 66 years (Interquartile range [IQR]: 59-72). Nearly half (49%) of the patients were female, and a high percentage (84%) of the patients were either current or former smokers. Adenocarcinoma was the most common histology, accounting for 72% of the cases, and 78% of patients had stage IV disease. The majority (92%) of patients received PD-1/PD-L1 inhibitor monotherapy. Among the 1,327 patients overall, 175 (13%) experienced ICI cessation due to irAEs, with 61 (4.6%) experiencing ICI cessation due to irAEs less than three months from the start of therapy (defined as early ICI cessation). The median time to ICI cessation due to irAEs was 6.8 (1.9 - 12.7) months.

**Table 1.**
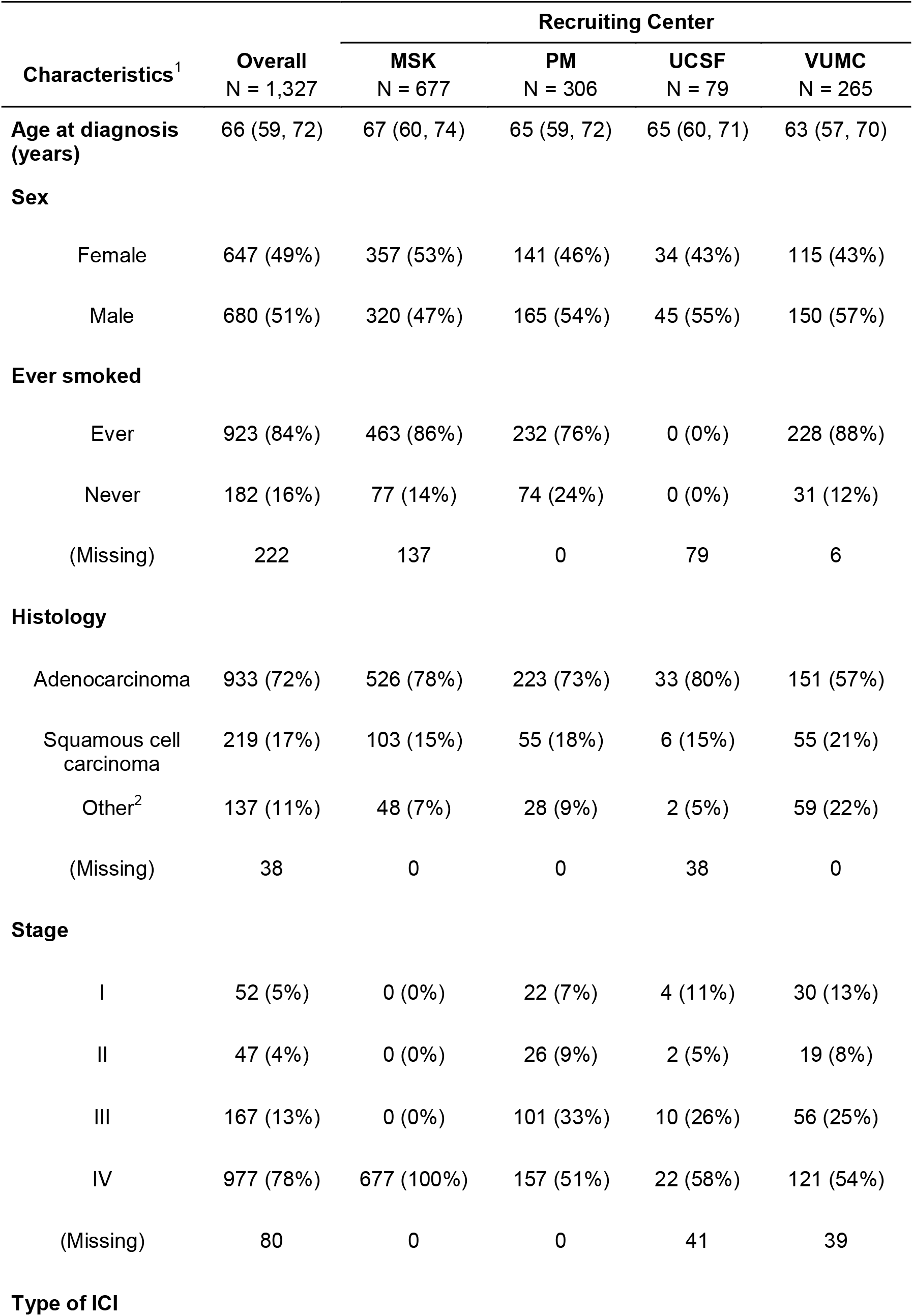
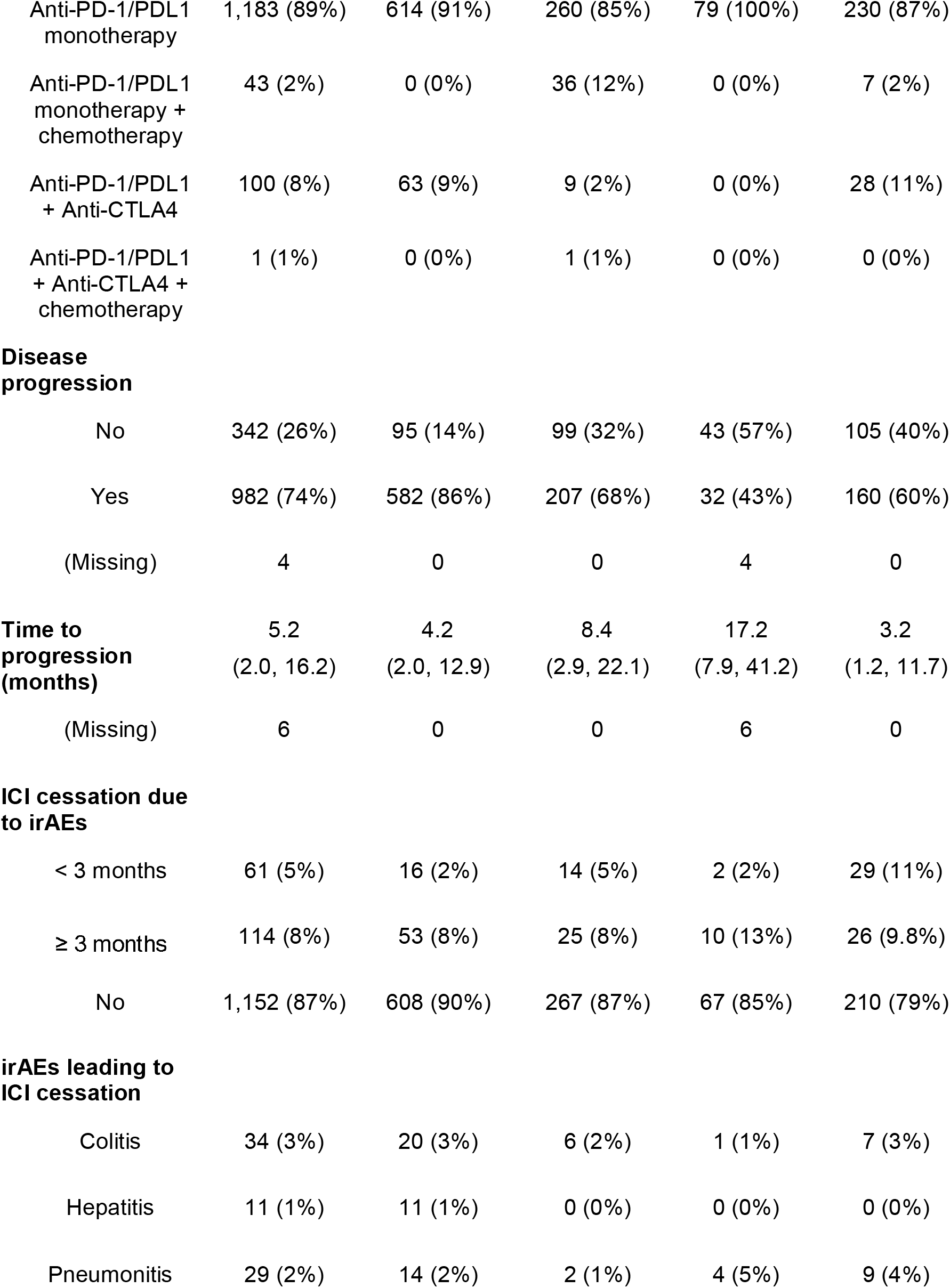
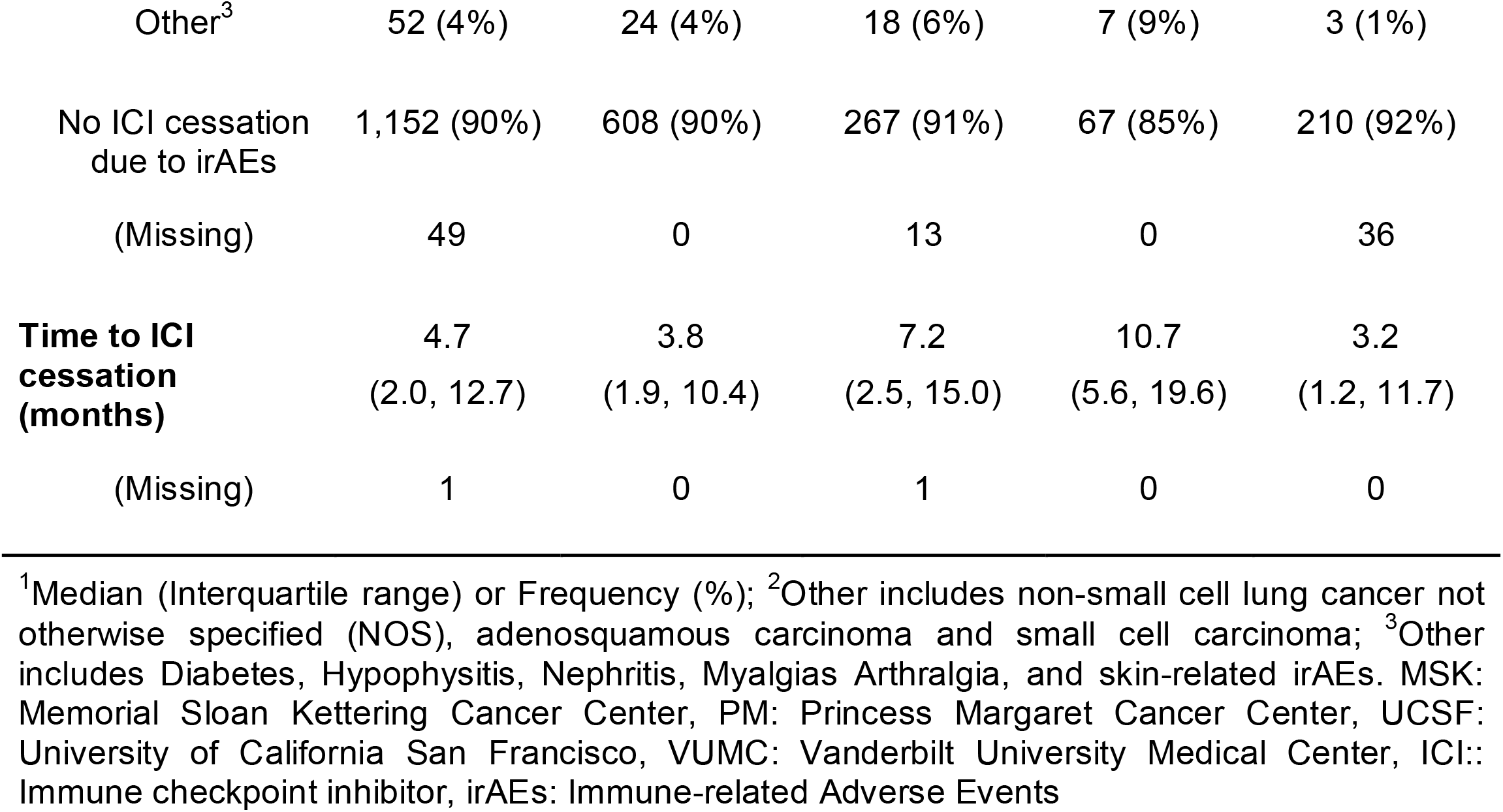
Patient characteristics in GeRI cohort: overall and by recruiting center.

### PRS_AD_ is associated with early ICI cessation due to irAEs

PRS_AD_ was associated with ICI cessation due to irAEs (HR per SD = 1.17, 95% CI = 1.01 - 1.36, P = 0.04). Individuals with high genetic risk (top 20th percentile of PRS_AD_) had an HR of 1.48 (95% CI = 1.05 - 2.09, P = 0.03). Individuals in the top 20th percentile of the PRS_AD_ had an increased cumulative events of ICI cessation due to irAEs, particularly early ICI cessation (7.2% early discontinuation among high-risk, 3.9% early discontinuation among low-risk by 3 months, log-rank P = 0.02) (Figure 2). Subsequently, we performed a competing risk model and observed a sub-distribution HR of 1.52 (95% CI = 1.08 - 2.14, P = 0.02). The association between PRS_AD_ and ICI discontinuation due to irAEs was stronger in individuals who had early ICI cessation due to irAEs (HR per SD = 1.38, 95% CI = 1.08 - 1.78, P = 0.01). We did not observe a significant association between PRS_AD_ and late ICI cessation due to irAEs (Table 2).

**Table 2:** Performance of polygenic risk score of autoimmune disease (standardized to have mean 0 and standard deviation of 1) on overall, early and late time to immune checkpoint inhibitor (ICI) therapy cessation due to immune-related adverse events (irAEs) in the GeRI cohort.

| ICI cessation due to irAEs | HR<sup>1</sup> | 95% CI | P |
| --- | --- | --- | --- |
| **Overall** |  |  |  |
| Continuous (per SD) | 1.17 | 1.01-1.36 | 0.04 |
| >80<sup>th</sup> percentile | 1.48 | 1.05-2.09 | 0.03 |
| **Early<sup>2</sup>** |  |  |  |
| Continuous (per SD) | 1.38 | 1.08-1.78 | 0.01 |
| >80<sup>th</sup> percentile | 1.80 | 1.03-3.14 | 0.04 |
| **Late<sup>2</sup>** |  |  |  |
| Continuous (per SD) | 1.08 | 0.89-1.31 | 0.43 |
| >80<sup>th</sup> percentile | 1.34 | 0.86-2.11 | 0.20 |

<sup>1</sup> All models are adjusted for age at diagnosis, sex, type of ICI therapy, lung cancer histology, recruiting center, and 5 principal components. <sup>2</sup> Early ICI cessation is defined as ICI cessation due to irAEs within 3 months of ICI therapy initiation, whereas late ICI cessation is defined as discontinuation of ICI therapy after 3 months from start of the ICI therapy

**Figure 2:**
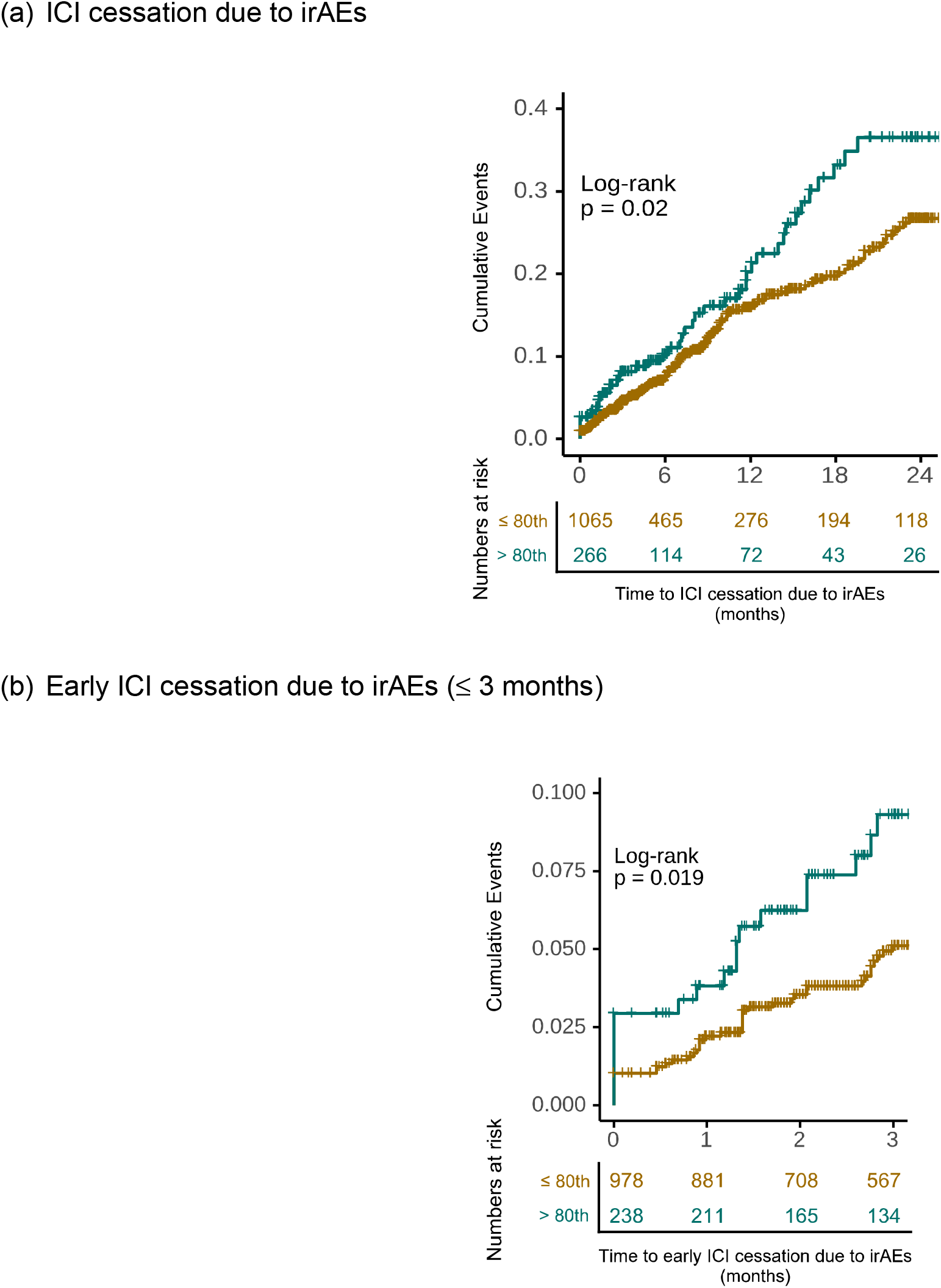
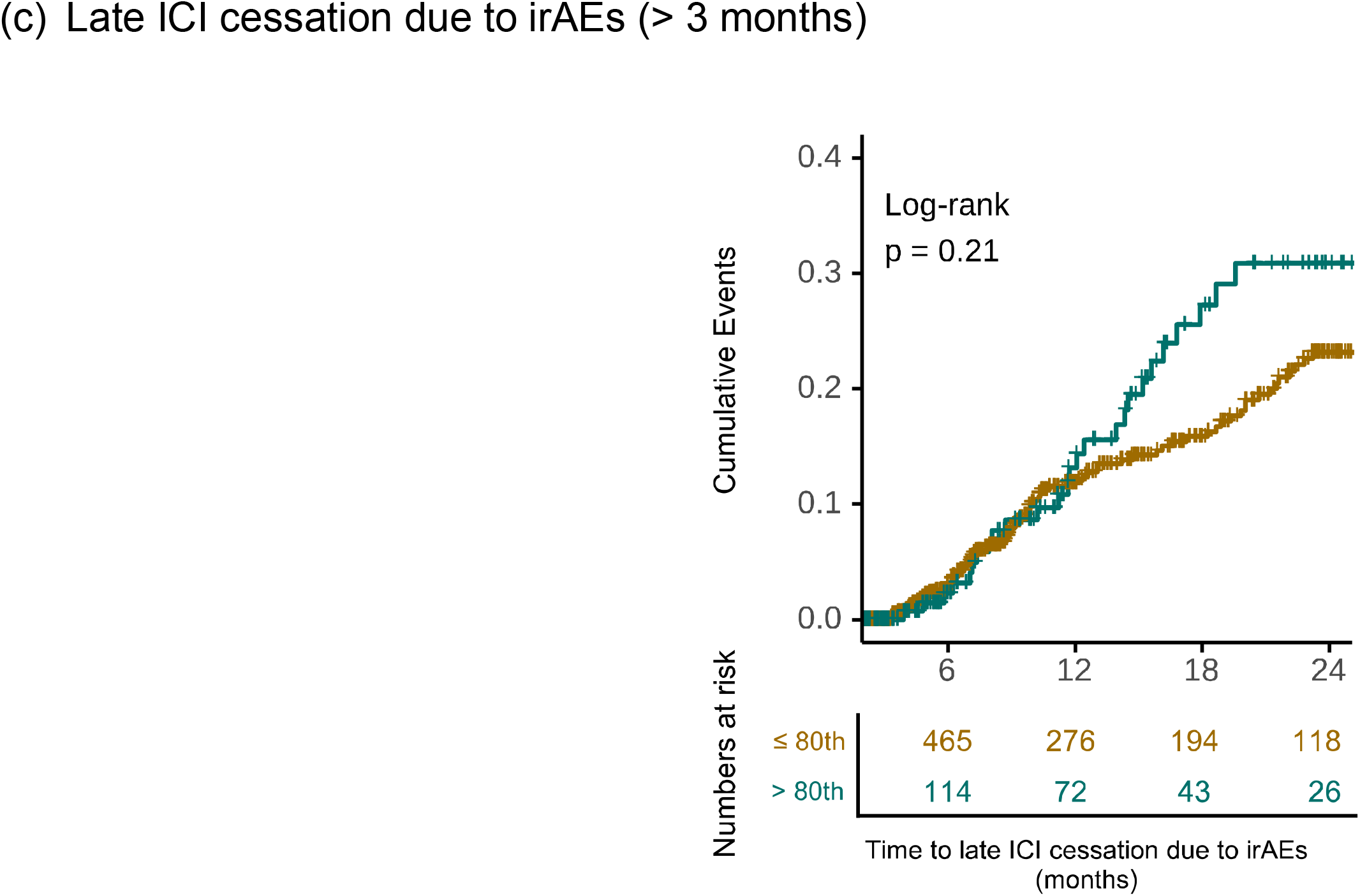
Cumulative incidence curves for (a) discontinuation of immune checkpoint inhibitor (ICI) therapy due to irAEs, (b) early discontinuation of ICI therapy due to irAEs, and (c) late discontinuation of ICI therapy due to irAEs across categories of polygenic risk score of autoimmune disease in the GeRI cohort. High genetic risk is defined as individuals in the >80^th^ percentile (top 20th percentile), whereas low/moderate genetic risk is defined as individuals in ≤80^th^ percentile. The p-values included on each plot are the results of a log-rank test for the difference between the curves (two-sided).

Additionally, we examined the association between PRS_AD_ and each irAE subtype leading to ICI discontinuation using a logistic regression model. Information regarding the type of irAE leading to discontinuation was available for 126 patients. Among 126 patients who had discontinuation of ICI therapy due to irAEs, the top irAEs were colitis (19.3%), pneumonitis (16.4%), and hepatitis (6.25%). Supplementary Table 1 reports the association for each irAE subtype. We observed a statistically significant association between PRS_AD_ and ICI cessation due to hepatitis irAE (OR per SD = 2.40, 95% CI = 1.23 - 4.93, P = 0.01).

### Association between PRS_AD_ and ICI cessation is stronger in individuals who received combination therapy

To characterize the differential association between PRS_AD_ and ICI cessation, we performed stratified analyses by type of ICI therapy and disease stage, (Table 3). Patients who received a combination of PD-1/PD-L1 inhibitors and CTLA4 inhibitors showed an OR per SD of 1.86 (95% CI = 1.08 - 3.51, P = 0.04), whereas, patients who received PD-1/PD-L1 monotherapy had an OR per SD of 1.22 (95% CI = 1.02 - 1.45, P = 0.03). In particular, early ICI discontinuation for irAEs occurred in 4 of the 19 patients with high PRS_AD_ genetic risk who received combination PD-1/PD-L1 inhibitors and CTLA4 inhibitors, whereas among 82 low PRS_AD_ genetic risk patients who received combination PD-1/PD-L1 inhibitors and CTLA4 inhibitors, 5 had early ICI discontinuation. When stratified by lung cancer stage, we found an OR per SD of 1.37 (95% CI = 1.11 - 1.69, P = 0.003) for patients with stage IV non-small cell lung cancer (NSCLC) (Table 3).

**Table 3:** Stratified logistic regression analyses assessing the association between polygenic risk score of autoimmune disease and immune checkpoint inhibitor cessation due to immune-related adverse events in GeRI cohort.

| Variable | N | OR per SD<sup>1</sup> | 95% CI | P |
| --- | --- | --- | --- | --- |
| **Type of ICI therapy<sup>2</sup>** |  |  |  |  |
| Anti- PD-1/PD-L1 monotherapy | 1,230 | 1.22 | 1.02 – 1.45 | 0.03 |
| Anti PD-1/PD-L1 inhibitor + Anti-CTLA4 inhibitor | 101 | 1.86 | 1.08 – 3.51 | 0.04 |
| **Stage<sup>3</sup>** |  |  |  |  |
| I | 56 | 1.87 | 0.77 – 5.19 | 0.18 |
| II | 47 | 1.19 | 0.46 – 3.13 | 0.72 |
| III | 167 | 1.10 | 0.72 – 1.64 | 0.66 |
| IV | 977 | 1.37 | 1.11 – 1.69 | 0.003 |

<sup>1</sup> PRS was standardized to have a mean of 0 and standard deviation of 1. <sup>2</sup> Models are adjusted for age at diagnosis, sex, lung cancer histology, recruiting center, and 5 principal components. ICI: Immune checkpoint inhibitor therapy, OR: Odds ratio, SD: Standard deviation, CI: Confidence interval

## Discussion

Here, using a large, multi-center, clinically annotated cohort of patients with lung cancer treated with ICIs, we demonstrate that a genetic polygenic risk score for autoimmune disease predicts the cessation of ICIs due to irAEs. Our results continue to build on a body of recent work^25–28^ describing the association of germline genetic variation on the risk of irAE development in patients with solid tumors treated with ICIs, and represent a further proof of principle that continued investigation of germline predictors of ICI toxicity is warranted.

Our results suggest that PRS_AD_ may broadly predict generalized severe irAEs. Given that low-grade irAEs may, in fact, be associated with improved clinical activity of ICIs, which has been demonstrated in multiple tumor types, including NSCLC^15,31^ and melanoma^31^, understanding predictors of high-grade irAEs, in particular, is critical to further assist with risk stratification for patients who are candidates for ICI therapy. Although our use of ICI cessation as a proxy for severe irAEs was a limitation of our approach, it is notable that PRS_AD_ more strongly associated with early ICI discontinuation due to irAEs and did not specifically associate with late discontinuation. We believe these results generally support the biological underpinnings of PRS_AD_ and severe irAEs, as in routine practice, early ICI discontinuation may be more likely to occur in patients with higher grade toxicity, whereas later in the course of treatment, clinicians may be more likely to discontinue irAEs for lower grade toxicity after a prolonged period of disease control. Correspondingly, in our study, among the patients in which grade of index irAE leading to ICI discontinuation was available, early ICI cessation was more frequently experienced for severe (G3+) irAEs compared to late cessation. These data overall suggest that the association of PRS_AD_ with early ICI cessation is clinically meaningful, and suggest that the biological underpinnings of PRS_AD_ with severity of ICI toxicity warrant further study.

In addition, the enriched association of PRS_AD_ with ICI discontinuation among patients receiving combination anti-PD-1/PD-L1 and anti-CTLA4 therapy suggests that PRS_AD_ may have particular utility in risk stratification in situations where the risk of ICI toxicity is highest^4,16^. Currently, for multiple indications across solid tumor oncology, including advanced melanoma^4,5^, renal cell carcinoma^8,32–34^, hepatocellular carcinoma^10,35^ and others, clinical equipoise exists around the decision to treat with either combination ICI therapy or ICI monotherapy with or without other therapies. If prospectively validated as a predictive marker of severe ICI toxicity and early cessation, PRS_AD_ may emerge as a useful adjunctive tool to help guide treatment decisions in these and related scenarios. In addition, given the emerging landscape of clinical trials investigating the use of prophylactic immunosuppressants to help maximize exposure to combination ICI therapy in advanced melanoma and other solid tumors^36,37^, individuals with high PRS_AD_ genetic risk may represent a subpopulation of patients who could particularly benefit from such approaches. Finally, an a priori understanding of the risk of development of severe irAEs could potentially assist clinicians with determining whether severe adverse events during the course of ICI treatment may be more or less likely to be immune-related, including irAEs such as pneumonitis, which can often be difficult to distinguish from alternative non-irAE etiologies clinically.

Despite the association of PRS_AD_ with early ICI discontinuation for irAEs demonstrated in our study, however, it is likely that additional features also contribute to the risk of severe irAE development in patients treated with ICIs. In addition to refining predictive germline genetic signatures, future efforts could include the development of multi-modal platforms incorporating tumor-intrinsic features^22^, host T cell^22^ or B cell repertoires^24^, features of host-microbiome^23^ and other components, which have been suggested to additionally predict for irAE development. This being said, the declining cost and turnaround time of germline sequencing may suggest that PRS_AD_ could be more easily incorporated into routine practice if prospectively validated as a marker of severe ICI toxicity.

Our study has several limitations. First, a small subset of patients included in our study lacked lung cancer staging information, limiting our ability to fully describe the impact of our analyses across different disease stages. Furthermore, the absence of specific information regarding the grade of irAEs leading to ICI discontinuation in most patients limits our power to assess the impact of severity of toxicities on patient outcomes. Despite these limitations, it is important to recognize that our study stands as one of the largest studies of germline genetics and irAEs in patients treated with ICIs, providing valuable insights into predictors of toxicity in this patient population.

In summary, we describe the utility of a germline polygenic risk score for autoimmune disease to help identify patients at high risk for early ICI cessation due to severe irAEs. Future efforts to continue to refine germline polygenic risk scores to predict severe ICI toxicity could further develop our understanding of irAE pathogenesis and assist with treatment decisions, especially in clinical scenarios in which the risk of severe irAE development is high.

## Data Availability

All data produced in the present study are available upon reasonable request to the authors

## Funding

This work was supported by the National Institutes of Health R01-CA227466 and K24-CA169004 to E.Ziv; C.M.Lovly was supported in part by NIH NCI UG1CA233259, P01CA129243, and P30CA068485; R.Thummalapalli was supported by T32-CA009207; The Lusi Wong Fund, Posluns Fund, Alan Brown Chair in Molecular Genomics, Princess Margaret Cancer Foundation were awarded to G. Liu for this work; M.C. Aldrich was supported in part by R01-CA227466, U01CA253560, R01CA251758 and the Vanderbilt Institute for Clinical and Translational Research (UL1TR002243); Z. Quandt was supported NIDDK DiabDocs K12DK133995 and a Larry L Hillblom Foundation Start-Up Grant; A.J.Schoenfeld, D.Faleck were supported by the Memorial Sloan Kettering Cancer Center Support Grant/Core (P30-CA008748), the Druckenmiller Center for Lung Cancer Research at Memorial Sloan Kettering Cancer Center. The samples and/or dataset(s) used for the analyses described were obtained from Vanderbilt University Medical Center’s BioVU which is supported by numerous sources: institutional funding, private agencies, and federal grants. These include the NIH-funded Shared Instrumentation Grant S10OD017985 and S10RR025141; and CTSA grants UL1TR002243, UL1TR000445, and UL1RR024975. Genomic data are also supported by investigator-led projects that include U01HG004798, R01NS032830, RC2GM092618, P50GM115305, U01HG006378, U19HL065962, R01HD074711; and additional funding sources listed at https://victr.vumc.org/biovu-funding/.

## Acknowledgments

Princess Margaret Lung Group: Natasha B. Leighl, Penelope A. Bradbury, Frances A. Shepherd, Adrian G. Sacher & Lawson Eng

## Notes

### Competing Interest Statement

The authors have declared no competing interest.

### Funding Statement

Funding This work was supported by the National Institutes of Health R01CA227466 to EZiv
CMLovly was supported in part by NIH NCI UG1CA233259 P01CA129243 and P30CA068485
RThummalapalli was supported by T32CA009207
The Lusi Wong Fund Posluns Fund Alan Brown Chair in Molecular Genomics Princess Margaret Cancer Foundation were awarded to G Liu for this work
MC Aldrich was supported in part by R01CA227466 U01CA253560 R01CA251758 and the Vanderbilt Institute for Clinical and Translational Research UL1TR002243
Z Quandt was supported NIDDK DiabDocs K12DK133995 and a Larry L Hillblom Foundation StartUp Grant
AJSchoenfeld was supported by the Memorial Sloan Kettering Cancer Center Support Grant/Core P30CA008748 the Druckenmiller Center for Lung Cancer Research at Memorial Sloan Kettering Cancer Center
The samples and/or datasets used for the analyses described were obtained from Vanderbilt University Medical Center's BioVU which is supported by numerous sources: institutional funding private agencies and federal grants These include the NIHfunded Shared Instrumentation Grant S10OD017985 and S10RR025141 and CTSA grants UL1TR002243 UL1TR000445 and UL1RR024975 Genomic data are also supported by investigatorled projects that include U01HG004798 R01NS032830 RC2GM092618 P50GM115305 U01HG006378 U19HL065962 R01HD074711 and additional funding sources listed at https://victrvumcorg/biovufunding/
Princess Margaret Lung Group includes Natasha B Leighl Penelope A Bradbury Frances A Shepherd Adrian G Sacher & Lawson Eng

### Author Declarations

Ethics committee/IRB of University of California San Francisco gave ethical approval for this work Ethics committee/IRB of Vanderbilt University Medical Center gave ethical approval for this work Ethics committee/IRB of Memorial Sloan Kettering Cancer Center gave ethical approval for this work Ethics committee/IRB of Princess Margaret Cancer Center gave ethical approval for this work

