## Supplemental Materials for "Germline prediction of immune checkpoint inhibitor discontinuation for immune-related adverse events"

| **Supplementary Table 1: Association of polygenic risk score of autoimmune disease by irAE subtype among those who had ICI cessation due to irAEs in the GeRi cohort, using a logistic regression model.** | | | | |
| --- | --- | --- | --- | --- |
| irAE subtype | N^1^ | OR^2^ | 95% CI | *P* |
| Colitis | 34 | 0.90 | 0.63 – 1.28 | 0.57 |
| Pneumonitis | 29 | 1.31 | 0.87 – 1.97 | 0.20 |
| Hepatitis | 11 | 2.40 | 1.23 – 4.93 | 0.01 |
| Other^3^ | 52 | 1.27 | 0.94 - 1.72 | 0.12 |
| ^1^ Among 122 out of 176 who had information on irAE leading to ICI discontinuation. ^2^ Models adjusted for age at diagnosis, sex, type of ICI therapy, lung cancer histology, recruiting site, and 5 principal components. ^3^ Other include diabetes, hypophysitis, nephritis, myalgia, arthralgia, and skin-related irAEs. irAEs: immune-related adverse events, ICI: immune checkpoint inhibitor, OR: Odds ratio, CI: confidence interval. | | | | |
